# Dietary Patterns, Circulating Metabolome, and Risk of Type 2 Diabetes

**DOI:** 10.1101/2025.08.11.25333425

**Authors:** Huan Yun, Jie Hu, Vishal Sarsani, Xavier Loffree, Kai Luo, Buu Truong, Fenglei Wang, Magdalena Sevilla-Gonzalez, Deirdre K Tobias, Daniela Sotres-Alvarez, Jianwen Cai, Bharat Thyagarajan, Oana Zeleznik, Mercedes Sotos-Prieto, Robert D Burk, Yasmin Mossavar-Rahmani, Josiemer Mattei, Simin Liu, A. Heather Eliassen, Johanna W Lampe, Kathryn M. Rexrode, Clary B. Clish, Qi Sun, Eric Boerwinkle, Robert C. Kaplan, Walter C. Willet, JoAnn E. Manson, Bing Yu, Qibin Qi, Frank B. Hu, Liming Liang, Jun Li

**Affiliations:** Department of Epidemiology, Harvard T.H. Chan School of Public Health, Boston, MA, USA; Epidemiology and Center for Genomic Medicine and Department of Anesthesia, Critical Care and Pain Medicine, Massachusetts General Hospital and Harvard Medical School, Boston, MA, USA; Department of Statistics and Actuarial Science, University of Waterloo, ON, Canada; Department of Epidemiology and Population Health, Albert Einstein College of Medicine, Bronx, NY, USA; Department of Nutrition, Harvard T.H. Chan School of Public Health, Boston, MA, USA; Clinical and Translational Epidemiology Unit, Mongan Institute, Massachusetts General Hospital. Boston, Massachusetts, USA; Department of Biostatistics, Gillings School of Global Public Health, University of North Carolina at Chapel Hill, Chapel Hill, NC, USA; Department of Laboratory Medicine and Pathology, University of Minnesota, MN, MA; Channing Division of Network Medicine, Department of Medicine, Brigham and Women’s Hospital and Harvard Medical School, Boston, MA, USA; Department of Preventive Medicine and Public Health, School of Medicine, Autonomous University of Madrid, Madrid, Spain; Department of Environmental Health, Harvard T.H. Chan School of Public Health, Boston, MA, USA; Center for Global Cardiometabolic Health, Department of Epidemiology, Brown University, Providence, Rhode Island, USA; Public Health Sciences Division, Fred Hutchinson Cancer Center, Seattle, WA, USA; Department of Epidemiology, University of Washington, Seattle, WA, USA; Division of Women’s Health, Department of Medicine, Brigham and Women’s Hospital and Harvard Medical School, Boston, MA, USA; Broad Institute of MIT and Harvard, Boston, MA, USA; Human Genetics Center, School of Public Health, University of Texas Health Science Center at Houston, Houston, TX, USA; Human Genome Sequencing Center, Baylor College of Medicine, Houston, TX, USA; Division of Preventive Medicine, Department of Medicine, Brigham and Women’s Hospital and Harvard Medical School, Boston, MA, USA; Department of Biostatistics, Harvard T.H. Chan School of Public Health, Boston, MA, USA

## Abstract

Circulating metabolites may reflect biological homeostasis and have been linked to dietary intakes and human health, and may hold the promises to facilitate objective assessments of intakes and metabolic response to diets. Here, we integrated metabolomic, genetic, and metagenomic data from five longitudinal cohorts comprising 21,474 participants of diverse ethnic backgrounds, to develop metabolomic signatures for popular dietary patterns (i.e., three guideline-based diets, three plant-based diets, and two mechanism-based diets) and systematically investigated their clinical relevance. Applying machine-learning models in two deeply-phenotyped lifestyle validation studies, we identified eight metabolomic signatures (each included 37 to 66 metabolites) significantly correlated with their respective dietary pattern indices, consistently across multiple independent validation cohorts (r = 0.11–0.38; *P* < 8.06×10⁻⁹). These signatures included shared metabolites between diets (e.g., up to 67% among guideline-based diets, including hippuric and 3-indolepropionic acid), and metabolites unique to specific diets (e.g., N6,N6,N6-trimethyllysine to proinflammatory diet). In multivariable-adjusted analyses of 5 prospective cohorts (1,832 incident cases during up to 27 years of follow-up), the metabolomic signatures of healthful diets (i.e., Mediterranean and healthful plant-based diets) were associated with lower T2D risk (HR: 0.82–0.90; *P* < 3×10⁻⁶), while signatures for unhealthy diets (e.g., proinflammatory and hyperinsulinemia diets) were associated with higher T2D risk (HR: 1.23–1.26; *P* < 2×10⁻¹⁵); these associations were further supported by Mendelian randomization analysis incorporating genetic data. Finally, through genome-wide and taxa-wide associating analyses, we identified 15 genetic loci – including those involved in fatty acid and energy metabolism (e.g., *FADS1/2* and *CERS4*; *P* < 5×10^-8^), and 39 gut microbial species – including those relevant to butyric acid metabolism (e.g., *E. eligens* and *F. pranusnitzii*; FDR < 0.05), significantly associated with the metabolomic signatures of diets. Genetic variants and gut microbial diversity explained up to 19.1% and 10.6% of the variation in these signatures, respectively, underscoring a potential role of host genetics and gut microbiota in dietary metabolism. In conclusion, our study identified metabolomic signatures reflecting both intakes and individual metabolic response to various diets and are associated with future T2D risk. These signatures may facilitate individualized dietary assessments and risk stratification in future nutritional research.

## Introduction

Type 2 diabetes (T2D) imposes an enormous burden on the healthcare system and is projected to affect 1 in 8 adults by 2045. About 70% of T2D cases are attributable to suboptimal diet^1^ and potentially preventable by following a healthy dietary pattern at every life stage as recommended by the 2020-2025 Dietary Guidelines for Americans^2^. Several dietary patterns that emphasize the overall dietary quality, such as the Mediterranean diet (MED) and Dietary Approaches to Stop Hypertension (DASH), as well as those that prioritize high-quality plant-based foods (e.g., healthful plant-based diet), have been consistently associated with lower risk of T2D^3,4^. Recent studies examining the relationship between foods and mechanistic biomarkers linked to T2D, also found that dietary patterns that promote systemic inflammation and hyperinsulinemia were strongly associated with higher T2D risk, independent of other well-established dietary quality indices^5^. Despite this, the metabolic pathways linking these dietary patterns to T2D risk remain unclear, and the relationship between individual variations in metabolic responses to diets^6^ and T2D risk, is yet to be elucidated in order to better inform the development of effective dietary strategies.

The blood metabolome, composed of small molecules generated from biological processes, has the potential to reflect metabolic homeostasis as combined effect of the interplay among diet, genetics, gut microbiome, and other factors^7^. Prior studies have identified multiple metabolites associated with intake of various foods and dietary patterns^8–11^. Of note, in our previous study, we identified a multiple-metabolite profile (i.e., a metabolomic signature) of the Mediterranean diet that reflected both dietary intake and individual metabolic response, potentially influenced by genetic variants related to fatty acid and amino acid metabolism, and was associated with lower risk of cardiovascular disease (CVD) independent of dietary intake itself^12^. However, no study has systematically examined and compared metabolomic signatures for various dietary patterns and their associations with T2D risk, and how genetic variants and gut microbiota may affect metabolic responses to different dietary patterns are yet to be elucidated.

To fill these knowledge gaps, we systematically analyzed dietary, metabolomics, genomics, metagenomics, and clinical data from 5 U.S. prospective cohorts with diverse ethnic backgrounds, including the Nurses’ Health Study (NHS) and II, Health Professionals Follow-up Study (HPFS), Hispanic Community Health Study/Study of Latinos (HCHS/SOL), and Women’s Health Initiative (WHI). We identified, validated, and compared circulating metabolomic signatures for 8 dietary patterns, including 3 guideline-recommended diets, 3 plant-based diets, and 2 mechanism-driven diets, and their association with T2D risk. We further explored genetic variants and microbial species related to these dietary metabolomic signatures to provide novel insights into the roles of host genetic and gut microbiota in individual variability of dietary metabolism (**Figure 1**).

**Figure 1.**
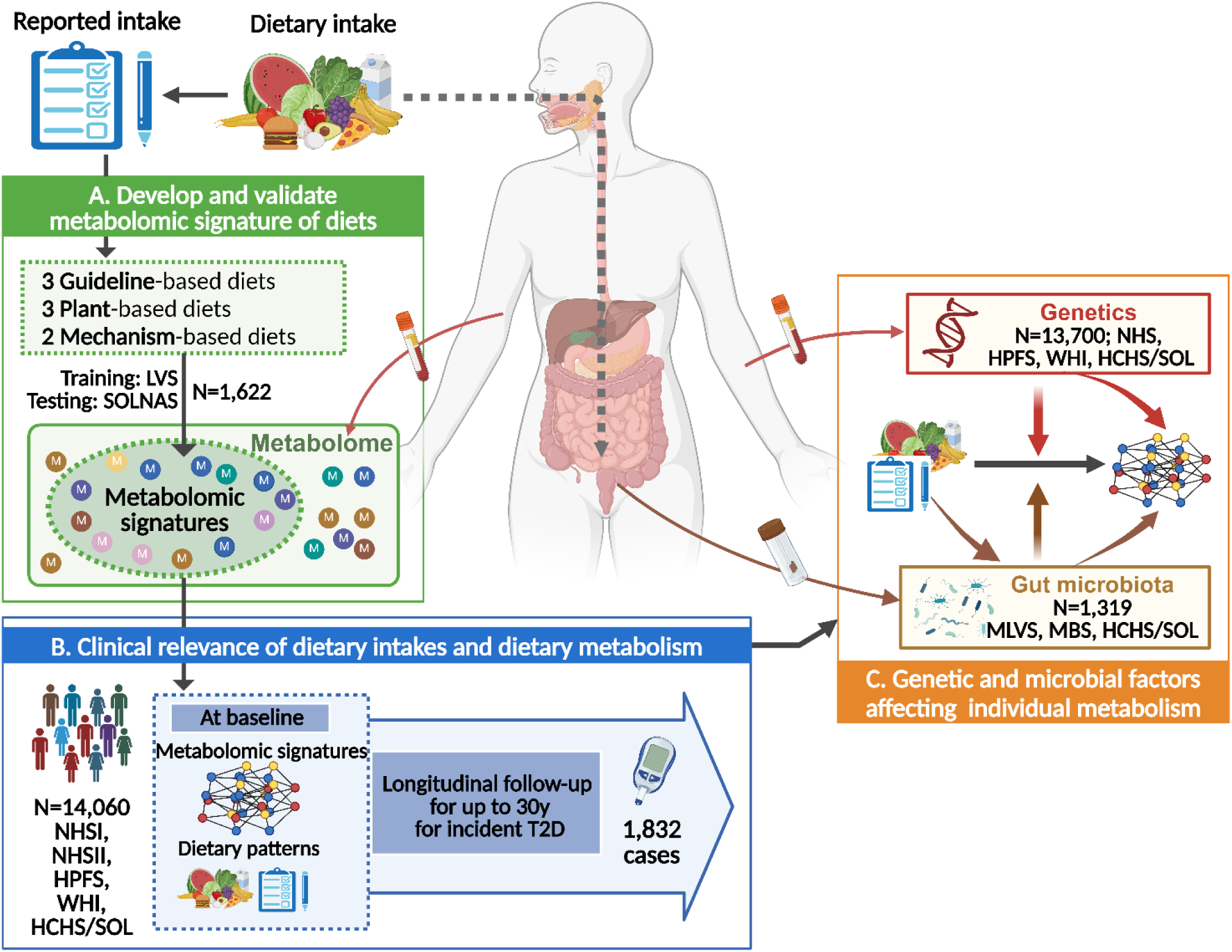
Overview of study design. **A.** We integrated diet and metabolome data to derive the metabolomic signature for each of 8 well-studied dietary patterns using LVS (n=1,206) as the training set and SOLNAS as the testing set (n=416). **B.** We applied the signatures from the elastic net model to five prospective cohorts, namely NHS, NHSII, HPFS, HCHS/SOL, and WHI and investigated the ability of metabolomic signatures of diet to predict risk of type 2 diabetes, followed by mediation analysis to explore to what extent the shared and unique metabolites across different dietary indices contribute to diet-diabetes associations. **C.** Within the sub-studies from NHS/HPFS and HCHS/SOL, we conducted a genome-wide association meta-analysis and microbiome-wide association analysis to examine to what degree genetics and gut microbiome explain the individual variation in dietary signatures; and explored the interaction between diet and genetics/microbiome on dietary metabolism. **Abbreviations:** HCHS/SOL, Hispanic Community Health Study/Study of Latinos; HPFS, Health Professionals Follow-up Study; LVS, Lifestyle Validation Study; MBS, Mind Body Study; NHS, Nurses’ Health Study; SOLNAS, Study of Latinos: Nutrition & Physical Activity Assessment Study; T2D, type 2 diabetes; WHI, Women’s Health Initiative.

## Methods

### Study populations

The NHS, NHSII, and HPFS are on-going prospective cohorts that started in 1976^13^, 1989^14^, and 1986^15^, respectively. At baseline, NHS enrolled 121,700 female nurses aged 30–55 years, NHSII enrolled 116,429 female nurses aged 25–42 years, and HPFS recruited 51,529 male professionals aged 40–75 years. Information on lifestyle, medical history, and health conditions was collected through self-administered questionnaires and updated biennially thereafter. Blood samples were collected in subsamples of NHS during 1989-1990 (n=32,826), NHSII during 1996-1999 (n=29,611), and HPFS during 1993-1995 (n=18,018). The plasma metabolome was profiled in several sub-studies (**Table S1**). For metabolomic signature development, we utilized dietary and metabolomic data from two sub-studies, the Men’s and Women’s Lifestyle Validation Studies (LVS, conducted between 2010-2013, n=1,206),^16,17^ in which dietary data were repeatedly collected over one year. For prospective analysis with T2D risk, we included 9,329 NHS/NHSII/HPFS participants with metabolomic data and were free of CVD, cancer, and T2D at the time of blood collection. To examine the role of gut microbiota in dietary metabolism, we utilized metabolomic and metagenomic data from the MLVS and the Mind body Study (MBS, a sub study within NHSII, N=202)^18^ (**Figure S1**). The study protocols were approved by the institutional review boards (IRB) of Harvard T.H. Chan School of Public Health and Brigham and Women’s Hospital, with participants’ consent implied by the return of the questionnaires.

The HCHS/SOL^19^ is a prospective community-based cohort launched between 2008 and 2011, enrolling 16,415 Hispanic/Latino adults aged 18–74 years from four U.S. sites. The first follow-up visit (Visit 2) was conducted between 2014 and 2017. The structured questionnaires were used to collect information on demographics, lifestyle, health status, family and medical histories, and medication use. Each visit included standardized clinical measurements and overnight fasting venous blood collection. First, we used 416 participants from the Nutrition and Physical Activity Assessment Study (SOLNAS)^20^ an ancillary study to HCHS/SOL to replicate dietary metabolomic signatures developed in LVS (**Figure S2**). In addition, 3,393 HCHS/SOL participants who had baseline blood metabolomics data, were included in the prospective analysis of T2D risk. Further, we used metabolomic and metagenomic data from 810 HCHS/SOL participants from the Gut Origins of Latino Diabetes (GOLD) ancillary study to analyze the role of gut microbiota. These study protocols were approved by the IRB at participating institutions, and all participants provided written informed consent.

The WHI^21^ is a nation-wide prospective study launched in 1991. At baseline, 68,132 women aged 50–79 years were enrolled either in Hormone Therapy Trials (WHI-HT), a Dietary Modification Trial, or a Calcium/Vitamin D Trial, while 93,676 women who did not participate in a clinical trial entered the Observational Study (WHI-OS). After the trial ended, participants were followed up through WHI Extension Studies (2005-2010, 2010-2015, and 2015-2020). A total of 1,338 participants, whose fasting plasma metabolome was profiled in a nested case-control study of coronary heart disease, were free of CVD, diabetes, and cancer at baseline, not included in the active arm of the Dietary Modification Trial, and were used as a replication study in our prospective analysis of T2D risk. The study was approved by the institutional review boards and all participants provided written informed consent. In this study, race categories as defined by NIH (American Indian or Alaska Native; Asian or Pacific Islander; Black or African American; Hispanic/Latino; White, Other) were collapsed, considering that race differed across studies in such a way.

### Assessment of diet, and calculation of dietary pattern indices

In the full cohort of NHS/HPFS, diet was assessed using a validated semi-quantitative food frequency questionnaire (FFQ)^22^ every 4 years, whereas in LVS, FFQ was administrated twice within a year. In the WHI, diet was assessed using a validated semi-quantitative FFQ, compatible to that used in the NHS/HPFS but tailored for application in multiethnic and geographically diverse populations^23^. In HCHS/SOL, diet was assessed by two 24-h dietary recalls at the first visit^20^. Our study focused on 8 dietary indices previously associated with T2D prevention; these include, an alternate MED (AMED) assessing adherence to a Mediterranean-style diet (9 components); the alternate healthy eating index (AHEI) assessing overall diet quality (11 components); DASH diet for hypertension prevention (8 components); three Plant-based Dietary Indices (PDI) that assess overall, healthful (hPDI), and unhealthful (uPDI) intakes of plant-based diets (18 components); as well as two data-driven mechanism-based diets, namely empirical dietary inflammatory pattern (EDIP) and empirical dietary index for hyperinsulinemia (EDIH; each with 18 components). Individual components and scoring criteria are disclosed in **Table S2**. Distributions of dietary indices and individual components in each study and characteristics according to tertile of these indices were presented in **Table S3** and **Table S4**, respectively.

### Metabolomics profiling and quality filtering

Circulating metabolomics in NHS/HPFS (including LVS and MBS), SOLNAS, and WHI were profiled using high-throughput liquid chromatography-mass spectrometry (LC-MS) at the Broad Institute of M.I.T and Harvard University (Cambridge, Massachusetts, USA)^24^. Two or more of methods were applied across different sub-studies: *(1)* the HILIC-pos that measured water-soluble metabolites including acylcarnitines, amino acids, peptides, and derivates; *(2)* a HILIC-neg method that measured sugar, purines and pyrimidines; *(3)* a C8-pos method measuring diverse species of lipids; and *(4)* a C18-neg method measuring free and/or oxidized fatty acids and bile acids. In the main cohort of HCHS/SOL, serum metabolome was profiled using the LC-MS DiscoveryHD4 Platform at the Metabolon Inc. (Durham, NC)^25^.

In each study set, we excluded metabolites with a detection rate of <75%, or with an intraclass correlation coefficient <0.4 (in NHS/HPFS and LVS), or with coefficient of variation >25%, or with no inter-person variation and removed individuals with a missing rate >25%. After filtering, the remaining missingness for each metabolite was imputed with half of the minimum value of that metabolite. Data processing and quality control (QC) were conducted within each sub-study of metabolomic profiling within each cohort. To ensure the generalizability of metabolomic signatures of diets, we primarily used 122 quality-controlled metabolites overlapping between the Broad Institute Platform (that used in LVS) and Metabolon Platform to develop metabolomic signatures. All quality-controlled metabolites (n=286) from the Broad Institute Platform were used in sensitivity analyses (**Figure S3)**.

### Genome-wide genotyping and processing

Genotyping was conducted using six different arrays (Affymetrix 6.0, Illumina 317k, Illumina 550k, Illumina 610k, Illumina 660w, and Illumina OmniExpress) across sub-studies within NHS/HPFS^26^; an Illumina custom array in HCHS/SOL^27^; and three arrays (Affymetrix 6.0, Illumina OmniExpress, and Illumina OmniQuad) in WHI^28^. Imputation was carried out utilizing the Trans-Omics for Precision Medicine (TOPMED) in NHS/HPFS, and 1000 Genomes Project phase 3 in HCHS/SOL^29^ and WHI. We filtered genetic variants with a minor allele frequency <0.01 and imputation quality R^2^<0.8, retaining 8,689,500 variants in NHS/HPFS, 13,047,680 in HCHS/SOL, and 11,078,977 in WHI for genome-wide association analyses.

### Metagenomic sequencing and taxonomic profiling

In MLVS and MBS, fecal DNA was extracted using Nextera XT DNA Library Preparation Kit and sequenced by Illumina HiSeq-end shotgun sequencing^30^. QC was conducted following HMP protocol with KneadData and taxonomic features were profiled using MetaPhlAn2^31^. Fecal DNA in the GOLD was sequenced using a shallow-coverage shotgun sequencing protocol^29^. We excluded taxonomic features with a relative abundance < 0.01% in over 10% of all samples, leaving 151 species in MLVS, 153 in MBS, and 204 in HCHS/SOL for microbial analyses.

### Ascertainment of incident type 2 diabetes

In NHS/HPFS, participants who reported diabetes through biennial questionnaire were further validated by a supplemental questionnaire to confirm the diagnosis (symptoms, medications, diagnosis test). T2D was defined according to the criteria from the National Diabetes Data Group before 1998; the criterion for fasting glucose was changed to ≥126 mg/dL (7.0 mmol/L) in 1998, and glycated hemoglobin (HbA_1c_) ≥6.5% was further added in the criteria in 2010^5^. In HCHS/SOL, T2D was defined as fasting time >8 hours and fasting glucose ≥7.0 mmol/L (126 mg/dL), fasting ≤8 hours and non-fasting glucose ≥11.1 mmol/L (200 mg/dL), 2-hour post-oral glucose tolerance test glucose ≥11.1 mmol/L (200 mg/dL), HbA_1c_ ≥6.5%, self-report of treatment with antidiabetic medications, or self-reported physician-diagnosed diabetes^29^. In WHI, T2D was defined as self-report of physician-diagnosed diabetes treated with oral medication or insulin; the accuracy of the diagnosis has been validated by using medication and laboratory data^23^.

### Statistical analysis

To develop the metabolomic signatures for dietary patterns, we first standardized all metabolites to a mean of 0 and standard deviation (SD) of 1. We fitted elastic net regression model to regress each of the dietary indices on 122 quality-controlled metabolites (overlapping between the Broad Institute and Metabolon) in the training set (i.e., LVS). The derived model for each dietary pattern was then applied to a testing set (i.e., SOLNAS) to calculate the metabolomic signature for cross-population replication. The metabolomic signature was computed as the weighted sum of the selected metabolites, with weights equal to the coefficients derived from the model. Further, we applied the models to NHS/HPFS (in sub-studies that contain >60% metabolites needed; **Table S5**), HCHS/SOL, and WHI participants with metabolomic data for additional validation. In the training set, to calculate the unbiased metabolomic signatures and avoid overfitting, we employed a leave-one-out approach (LOOCV), in which each person’s signature score was derived from a model trained on the remaining participants. Pearson correlations were calculated between dietary indices and their corresponding metabolomic signatures to evaluate model performance. Several sensitivity analyses were conducted, including comparing the model performance across different metabolomic profiling sub-studies and disease status; deriving the metabolomic signatures using all 286 quality-controlled metabolites measured by the Broad Institute Platform; and employing 9 other machine-learning models using physics-informed machine learning (PiML).

We estimated the associations of dietary indices and metabolomic signatures with incident T2D using Cox regression in NHS/HPFS and WHI, and Poisson regression in HCHS/SOL. Model was adjusted for age, sex, self-reported race, original sub-studies (not in HCHS/SOL), the case-control status in the original sub-study (not in HCHS/SOL), smoking, alcohol consumption, physical activity in metabolic equivalent of energy (MET) per week, family history of diabetes, hypertension, dyslipidemia, lipid-lowering medications, aspirin (only in NHS/HPFS), total energy intake, and BMI (continuous). In sensitivity analyses, we further adjusted blood lipids and/or estimated glomerular filtration rate (eGFR) in separate models. We applied a mediation analysis, implemented in R package CMAverse^32^, to explore the degree to which the metabolomic signatures explained the associations between the corresponding dietary indices and T2D risk. We secondarily analyzed the associations of individual metabolites with dietary indices (and food/nutrient components) using multivariable linear regression, and with T2D risk using Cox or log-Poisson regressions, to explore key mediating metabolites.

To identify genetic determinants of dietary metabolomic signatures, we conducted genome-wide association study (GWAS) in the NHS/HPFS (n=7,288), HCHS/SOL (n=5,140), and WHI (n=1,257) using REGENIE software (v2.0.2)^33^. Inverse-normal transformed metabolomic signatures were first regressed on age, sex, top principal components, and other study-specific covariates, and model residuals were then used in genetic association analysis, regressed on genetic variants (model 1), or genetic variants, the corresponding dietary pattern index, and the interactions between genetic variants and the dietary index (model 2). Summary statistics of each signature from each cohort were combined using a fixed-effect meta-analysis using METAL^34^. Lead variants were identified using the Plink *-clump* function (*P* < 5×10^-8^ and r^2^<0.1 in a 1000kbp window). We applied linkage disequilibrium score regression to estimate SNP-based genetic heritability of metabolomic signatures and their genetic correlations with 25 T2D-related traits reflecting its pathophysiological mechanisms including insulin secretion, insulin resistance, post-load insulin/glucose response, obesity, fat deposition, blood lipids, and liver function. To infer the temporal relationship between the dietary metabolomic signatures and T2D risk, we performed a two-sample Mendelian randomization (MR) analysis using four methods (mode-based estimate of Hartwig, inverse variance weighted, weighted median, and MR-Egger), leveraging GWAS summary statistics for T2D from the DIAGRAM Consortium meta-analysis (80,154 cases and 853,816 controls)^35^.

Finally, to identify gut microbial species associated with dietary metabolomic signatures, we applied elastic net regression to regress each metabolomic signature on all microbial species that passed QC; the resulted microbial score (i.e., a weighted sum of selected species) was then used to estimate the variance (R^2^) explained by microbial composition in the metabolomic signatures using linear regression, adjusted for age, smoking, alcohol drinking, physical activity, antibiotic use, probiotics use, colonoscopy (not in HCHS/SOL), anti-acid medication use (not in HCHS/SOL), the Bristol stool chart (not in HCHS/SOL), and BMI. To avoid overfitting, the microbial scores were estimated using a 10-fold cross-validation approach, in which scores of 10% of participants were estimated using models derived from the rest 90%. Associations between selected taxa and metabolomic signatures were tested using multivariable-adjusted generalized linear regression, and feature with false discovery rate (FDR)< 0.05 was considered statistically significant. To explore the role of the gut microbiota in dietary metabolism, for each dietary pattern, we examined whether the gut microbial score mediated the association between the dietary index and the metabolomic signature using mediation analysis; and further explored whether the gut microbial score modified the associations between dietary index and the metabolomic signature using interaction analysis. The significance of the interaction term was estimated using a likelihood ratio test.

All analyses were performed using R (version 4.2.3) unless otherwise specified. Details on the abovementioned methods are provided in **Supplementary Methods**.

## Results

### Basic characteristics of the study participants

The dietary metabolomic signatures were primarily derived in 1,206 LVS participants (mean age=66.4 years; 61.8% women) and validated in 416 SOLNAS participants (mean age=46.0 years; 61.1% women). We then replicated the dietary metabolomic signatures in 6,741 women from NHS (mean age=57.0 years) and 3,408 from NHSII (mean age=44.6 years), 1,614 men from HPFS (mean age=63.9 years), 6,166 participants from HCHS/SOL (mean age=48.3 years; 60.0% women), and 1,923 women from WHI (mean age=67.2 years). Compared with participants from NHS/HPFS and WHI who were predominantly non-Hispanic White, Hispanic/Latino participants from SOLNAS and HCHS/SOL were more likely to be current smokers, had lower healthy dietary quality scores, but had higher levels of physical activity (**Table 1**).

**Table 1.**
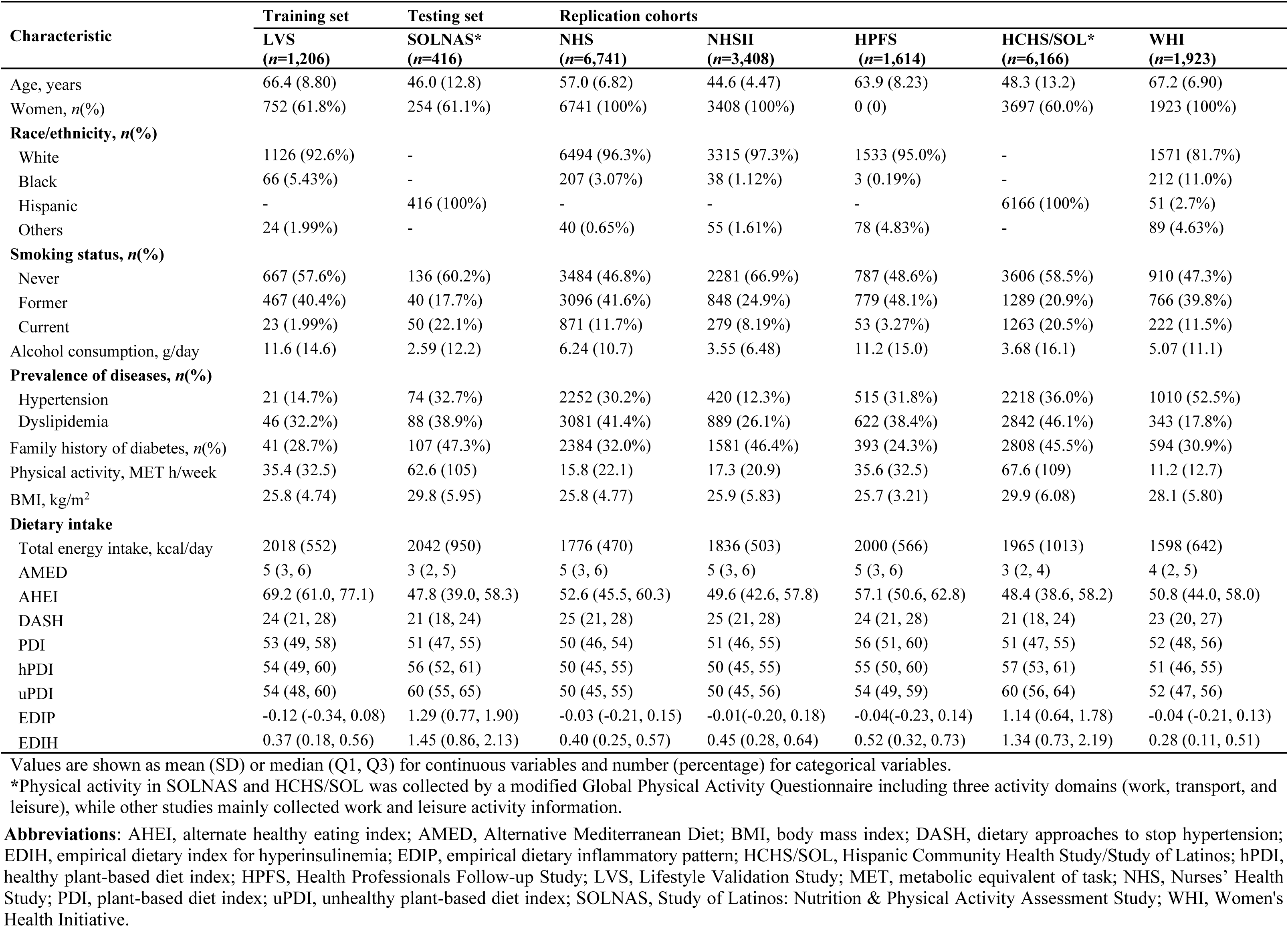
Characteristics of study participants.

| Characteristic | Training set | Testing set | Replication cohorts |  |  |  |  |
| --- | --- | --- | --- | --- | --- | --- | --- |
|  | LVS<br>(*n*=1,206) | SOLNAS*<br>(*n*=416) | NHS<br>(*n*=6,741) | NHSII<br>(*n*=3,408) | HPFS<br>(*n*=1,614) | HCHS/SOL*<br>(*n*=6,166) | WHI<br>(*n*=1,923) |
| Age, years | 66.4 (8.80) | 46.0 (12.8) | 57.0 (6.82) | 44.6 (4.47) | 63.9 (8.23) | 48.3 (13.2) | 67.2 (6.90) |
| Women, *n*(%) | 752 (61.8%) | 254 (61.1%) | 6741 (100%) | 3408 (100%) | 0 (0) | 3697 (60.0%) | 1923 (100%) |
| **Race/ethnicity, *n*(%)** |  |  |  |  |  |  |  |
| White | 1126 (92.6%) | - | 6494 (96.3%) | 3315 (97.3%) | 1533 (95.0%) | - | 1571 (81.7%) |
| Black | 66 (5.43%) | - | 207 (3.07%) | 38 (1.12%) | 3 (0.19%) | - | 212 (11.0%) |
| Hispanic | - | 416 (100%) | - | - | - | 6166 (100%) | 51 (2.7%) |
| Others | 24 (1.99%) | - | 40 (0.65%) | 55 (1.61%) | 78 (4.83%) | - | 89 (4.63%) |
| **Smoking status, *n*(%)** |  |  |  |  |  |  |  |
| Never | 667 (57.6%) | 136 (60.2%) | 3484 (46.8%) | 2281 (66.9%) | 787 (48.6%) | 3606 (58.5%) | 910 (47.3%) |
| Former | 467 (40.4%) | 40 (17.7%) | 3096 (41.6%) | 848 (24.9%) | 779 (48.1%) | 1289 (20.9%) | 766 (39.8%) |
| Current | 23 (1.99%) | 50 (22.1%) | 871 (11.7%) | 279 (8.19%) | 53 (3.27%) | 1263 (20.5%) | 222 (11.5%) |
| Alcohol consumption, g/day | 11.6 (14.6) | 2.59 (12.2) | 6.24 (10.7) | 3.55 (6.48) | 11.2 (15.0) | 3.68 (16.1) | 5.07 (11.1) |
| **Prevalence of diseases, *n*(%)** |  |  |  |  |  |  |  |
| Hypertension | 21 (14.7%) | 74 (32.7%) | 2252 (30.2%) | 420 (12.3%) | 515 (31.8%) | 2218 (36.0%) | 1010 (52.5%) |
| Dyslipidemia | 46 (32.2%) | 88 (38.9%) | 3081 (41.4%) | 889 (26.1%) | 622 (38.4%) | 2842 (46.1%) | 343 (17.8%) |
| Family history of diabetes, *n*(%) | 41 (28.7%) | 107 (47.3%) | 2384 (32.0%) | 1581 (46.4%) | 393 (24.3%) | 2808 (45.5%) | 594 (30.9%) |
| Physical activity, MET h/week | 35.4 (32.5) | 62.6 (105) | 15.8 (22.1) | 17.3 (20.9) | 35.6 (32.5) | 67.6 (109) | 11.2 (12.7) |
| BMI, kg/m<sup>2</sup> | 25.8 (4.74) | 29.8 (5.95) | 25.8 (4.77) | 25.9 (5.83) | 25.7 (3.21) | 29.9 (6.08) | 28.1 (5.80) |
| **Dietary intake** |  |  |  |  |  |  |  |
| Total energy intake, kcal/day | 2018 (552) | 2042 (950) | 1776 (470) | 1836 (503) | 2000 (566) | 1965 (1013) | 1598 (642) |
| AMED | 5 (3, 6) | 3 (2, 5) | 5 (3, 6) | 5 (3, 6) | 5 (3, 6) | 3 (2, 4) | 4 (2, 5) |
| AHEI | 69.2 (61.0, 77.1) | 47.8 (39.0, 58.3) | 52.6 (45.5, 60.3) | 49.6 (42.6, 57.8) | 57.1 (50.6, 62.8) | 48.4 (38.6, 58.2) | 50.8 (44.0, 58.0) |
| DASH | 24 (21, 28) | 21 (18, 24) | 25 (21, 28) | 25 (21, 28) | 24 (21, 28) | 21 (18, 24) | 23 (20, 27) |
| PDI | 53 (49, 58) | 51 (47, 55) | 50 (46, 54) | 51 (46, 55) | 56 (51, 60) | 51 (47, 55) | 52 (48, 56) |
| hPDI | 54 (49, 60) | 56 (52, 61) | 50 (45, 55) | 50 (45, 55) | 55 (50, 60) | 57 (53, 61) | 51 (46, 55) |
| uPDI | 54 (48, 60) | 60 (55, 65) | 50 (45, 55) | 50 (45, 56) | 54 (49, 59) | 60 (56, 64) | 52 (47, 56) |
| EDIP | -0.12 (-0.34, 0.08) | 1.29 (0.77, 1.90) | -0.03 (-0.21, 0.15) | -0.01(-0.20, 0.18) | -0.04(-0.23, 0.14) | 1.14 (0.64, 1.78) | -0.04 (-0.21, 0.13) |
| EDIH | 0.37 (0.18, 0.56) | 1.45 (0.86, 2.13) | 0.40 (0.25, 0.57) | 0.45 (0.28, 0.64) | 0.52 (0.32, 0.73) | 1.34 (0.73, 2.19) | 0.28 (0.11, 0.51) |

Values are shown as mean (SD) or median (Q1, Q3) for continuous variables and number (percentage) for categorical variables.
\*Physical activity in SOLNAS and HCHS/SOL was collected by a modified Global Physical Activity Questionnaire including three activity domains (work, transport, and leisure), while other studies mainly collected work and leisure activity information.
**Abbreviations:** AHEI, alternate healthy eating index; AMED, Alternative Mediterranean Diet; BMI, body mass index; DASH, dietary approaches to stop hypertension; EDIH, empirical dietary index for hyperinsulinemia; EDIP, empirical dietary inflammatory pattern; HCHS/SOL, Hispanic Community Health Study/Study of Latinos; hPDI, healthy plant-based diet index; HPFS, Health Professionals Follow-up Study; LVS, Lifestyle Validation Study; MET, metabolic equivalent of task; NHS, Nurses' Health Study; PDI, plant-based diet index; uPDI, unhealthy plant-based diet index; SOLNAS, Study of Latinos: Nutrition & Physical Activity Assessment Study; WHI, Women's Health Initiative.

### Discovery and validation of metabolomic signatures for eight dietary patterns

Among the 10 initially tested machine-learning models, elastic net regularization delivered the best prediction performance and was thus used as the main model to identify metabolomic signatures for the 8 dietary pattern indices (**Table S6a**). First, by regressing each of the 3 indices of guideline-based diets on the 122 metabolites shared between the Broad and Metabolon platforms in LVS, we identified a metabolomic signature comprising 48 metabolites for AMED, 61 metabolites for AHEI, and 55 metabolites for DASH (**Figure 2A**). Metabolites selected in the signatures encompass various biochemical categories that include lipids (27.8%-35.1%) and amino acids (34.9%-44.7%), with some cofactors and vitamins (2.3%-6.3%), nucleotides (1.5%-9.3%), and xenobiotics (6.5%-14.6%). The 3 guideline-based diets were moderately-to-highly correlated (*r*=0.45-0.81; **Figure S4**); consistent with these correlations, we found that 32 metabolites were shared across all 3 indices, whereas only 12.5%, 21.3%, and 9.1% metabolites were unique in the signatures of AMED, AHEI, and DASH, respectively (**Figure 3A** and **3D**). Applying similar models, we identified 3 metabolomic signatures, constituting 36, 38, and 43 metabolites, for overall, healthful, and unhealthful PDIs, respectively; and up to 41.8% of the metabolites were shared between the signatures of these PDIs (**Figure 3B** and **3E**). Finally, the metabolomic signatures for EDIP (66 metabolites) included 39 unique metabolites, compared to that of EDIH (37 metabolites) (**Figure 3C** and **3F**).

**Figure 2.**
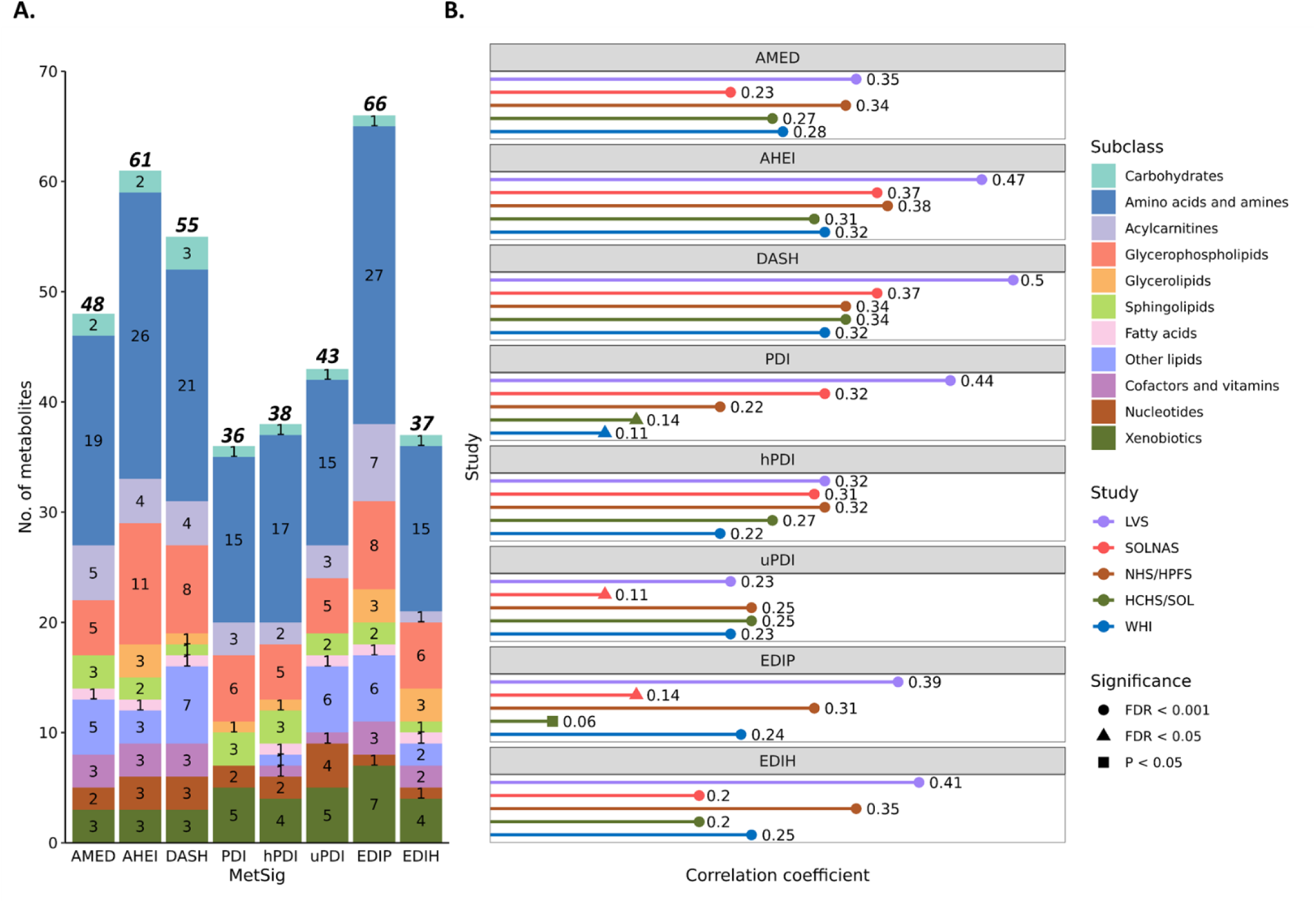
The metabolomic signatures for adherence to dietary patterns. **A.** Subclasses of metabolites constituting metabolomic signatures. **B.** The correlations between the metabolomic signatures and corresponding dietary pattern scores in each study. **Abbreviations**: AHEI, alternate healthy eating index; AMED, Alternative Mediterranean Diet; DASH, dietary approaches to stop hypertension; EDIH, empirical dietary index for hyperinsulinemia; EDIP, empirical dietary inflammatory pattern; FDR, false discovery rate; HCHS/SOL, Hispanic Community Health Study/Study of Latinos; hPDI, healthy plant-based diet index; HPFS, Health Professionals Follow-up Study; LVS, Lifestyle Validation Study; MetSig, metabolomic signature; NHS, Nurses’ Health Study; PDI, plant-based diet index; uPDI, unhealthy plant-based diet index; SOLNAS, Study of Latinos: Nutrition & Physical Activity Assessment Study; WHI, Women’s Health Initiative.

**Figure 3.**
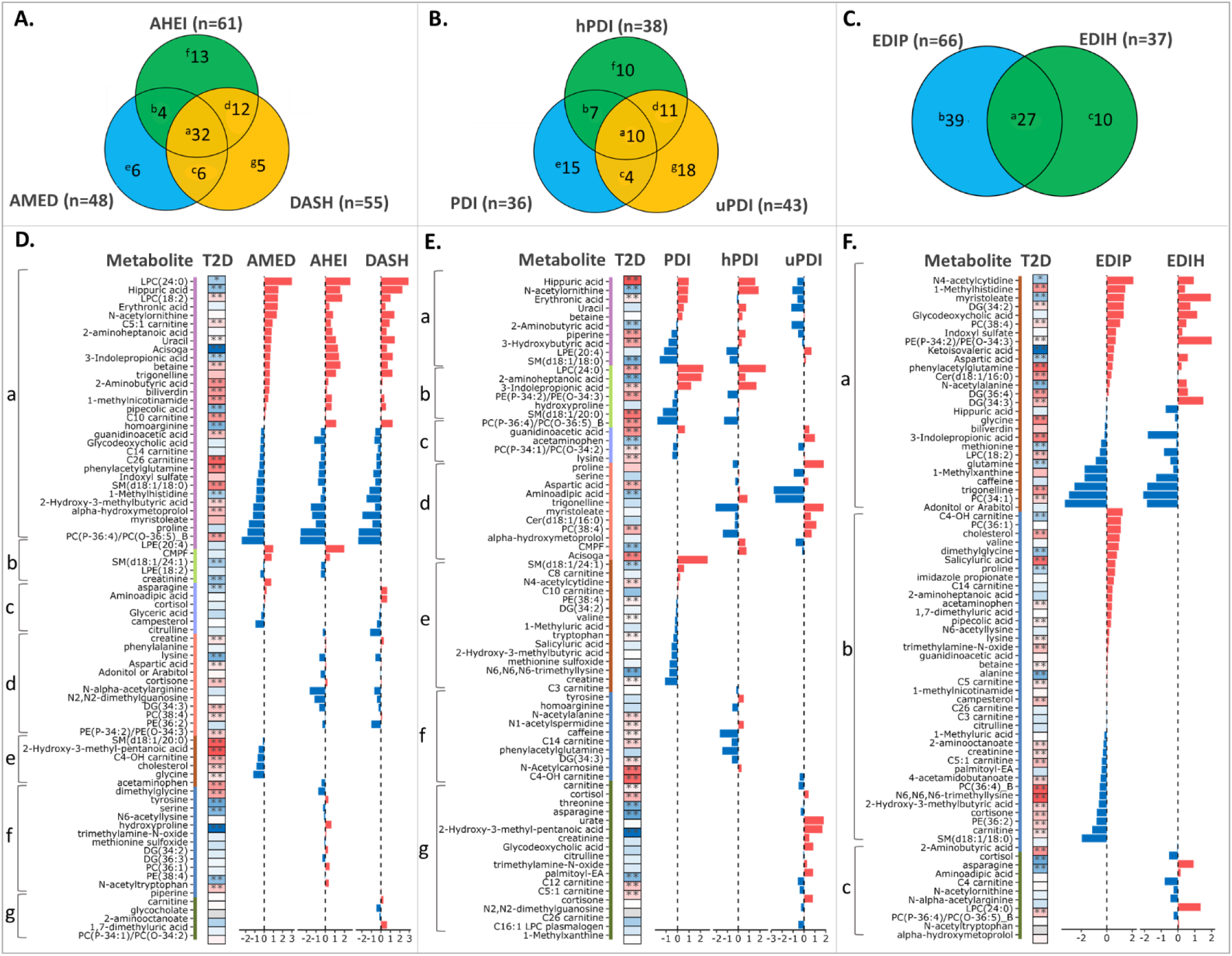
Similarity and differences across different dietary metabolomic signatures. **(A-C)** Venn plots displaying the overlapping metabolites among dietary guideline-recommended signatures (**A**), plant-based signatures (**B**), and mechanism-driven dietary signatures (**C**). **(D-F)** Presented from left to right are the metabolites’ characteristics, with each part corresponding to **A-C** (*see* color bar), the individual association with incident T2D (*see* heatmap; blue color indicates inverse association while red one denotes positive association), and coefficients (weights) in the signatures (*see* bar plot; blue color indicates negative coefficient while red one denotes positive coefficient). For the association with T2D incidence, we combined the results from NHS/HPFS, HCHS/SOL, and WHI. **, FDR corrected *P* < 0.01; *, FDR corrected *P* < 0.05. **Abbreviations:** AHEI, alternate healthy eating index; AMED, Alternative Mediterranean Diet; DASH, dietary approaches to stop hypertension; EDIH, empirical dietary index for hyperinsulinemia; EDIP, empirical dietary inflammatory pattern; HCHS/SOL, Hispanic Community Health Study/Study of Latinos; hPDI, healthy plant-based diet index; HPFS, Health Professionals Follow-up Study; NHS, Nurses’ Health Study; PDI, plant-based diet index; uPDI, unhealthy plant-based diet index; T2D, type 2 diabetes; WHI, Women’s Health Initiative.

Applying the aforementioned models to SOLNAS (the testing set), we verified that the metabolomic signatures were significantly correlated with corresponding dietary indices (AMED: *r*=0.23; AHEI and DASH: both *r*=0.37; PDI: *r*=0.32; hPDI: *r*=0.31; uPDI: *r*=0.11; EDIP: *r*=0.14; EDIH: *r*=0.20; all *P* < 3.01×10^-2^). The robust correlations were further replicated, when we applied these models to the main metabolomic cohorts of NHS/HPFS, HCHS/SOL, and WHI with diverse racial and ethnic backgrounds (total N=19,852; *r*=0.23-0.34 for AMED; *r*=0.31-0.38 for AHEI; *r*=0.32-0.37 for DASH; *r*=0.11-0.32 for PDI; *r*=0.22-0.32 for hPDI; *r*=0.11-0.25 for uPDI; *r*=0.24-0.31 for EDIP; *r*=0.20-0.35 for EDIH; all *P* < 3.83×10^-5^; **Figure 2B**). In the LVS (the training set), the correlations between the unbiased metabolomic signatures obtained using an LOOCV approach and the corresponding dietary indices were likewise, at a similar magnitude (*r*: 0.23-0.50; all *P* < 8.83×10^-17^). These correlations were consistent across sub-studies and disease status within cohorts (**Figure S5**). In sensitivity analysis, similar levels of performance of metabolomic signatures were noted when developing the signatures using all 286 quality-controlled metabolites from the Broad Institute metabolomic platforms (**Table S4b** and **Table S6b**; and **Figure S6** and **S7**).

When examining metabolites constituting the metabolomic signatures, we noted extensive associations with the corresponding dietary indices and specific food/nutrient components (**Figure S8**), suggesting the signatures may reflect the overall metabolomic profile related to various food components in each diet. For the three guideline-based diets, shared metabolites in their signatures also demonstrated relatively consistent weights that are partially due to their strong associations with shared major food components in these diets including vegetables, fruits, whole grains, and red meat (**Figure S6** and **S9**).

### Diet, metabolomic signatures, and risk of type 2 diabetes

To demonstrate the clinical relevance of the metabolomic signatures of diets, we examined their associations with incident T2D among 14,060 individuals free of diabetes, CVD, or cancer at baseline in NHS/HPFS, WHI, and HCHS/SOL. During up to 27 years follow-up, a total of 1,832 incident T2D cases were documented (**Table S7**). The results appeared to be generally consistent across cohorts. In the combined analysis of all 5 cohorts, after multivariable adjustment including BMI (continuous), higher metabolomic signatures for healthy diets, including AMED, AHEI, and DASH, PDI, and hPDI, were associated with lower T2D risk (relative risk [RR] and 95% confidence interval [CI] per SD increment in the signatures, were 0.87 [0.83-0.92], 0.82 [0.78-0.86], 0.89 [0.85-0.93], 0.81 [0.78-0.85], and 0.88 [0.83-0.92], respectively); whereas three higher metabolomic signatures for unhealthy diets, including uPDI, EDIP, and EDIH, were associated with higher T2D risk (RR and 95% CI, were 1.05 [1.00-1.10], 1.23 [1.17-1.29], and 1.26 [1.19-1.32], respectively). These associations were in similar directions, but stronger in magnitude, compared to the associations of their corresponding dietary indices; and remained significant, albeit slightly attenuated, after further adjusting for their corresponding dietary indices, except for uPDI (**Figure 4**). The results were generally consistent among individuals with or without obesity (*P*_interaction_ > 0.05; **Figure S10**) and did not materially change after adjusting for baseline blood lipids, or eGFR (*P*_heterogeneity_ > 0.05; **Table S8**). In the sensitivity analysis, when we developed the signatures based on all 286 metabolites from the Broad Institute platform, similar associations, albeit slightly weaker, were noted between the metabolomic signatures and T2D risk (**Table S9**).

**Figure 4.**
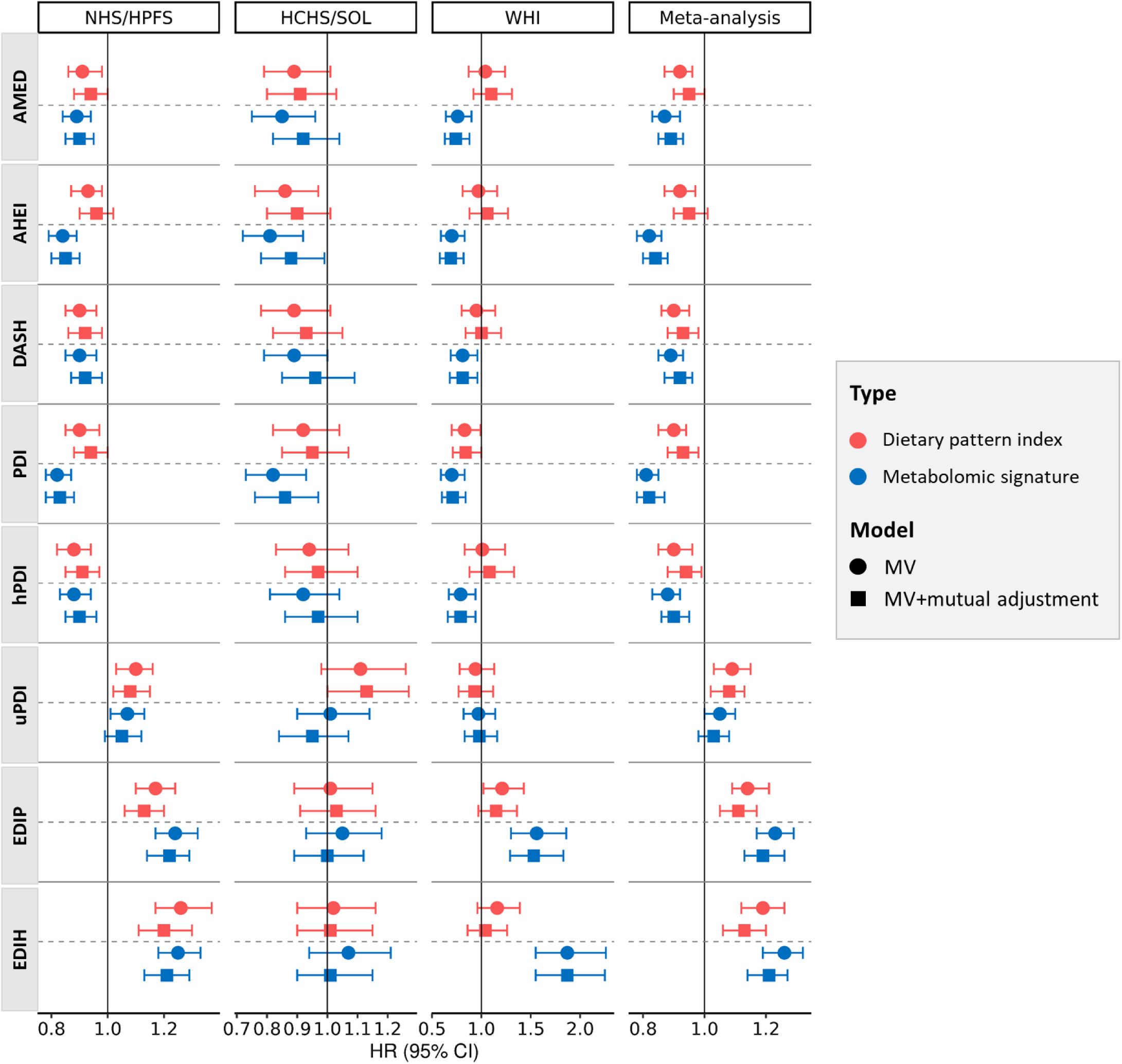
Associations of dietary pattern indices and metabolomic signatures with risk of type 2 diabetes. For the association with type 2 diabetes, we used Cox regression in NHS/HPFS and WHI and Poisson regression in HCHS/SOL. Multivariable (MV) adjusted model included age, sex, race/ethnicity, original sub-studies (not in HCHS/SOL), the case-control status in the original sub-study (not in HCHS/SOL), smoking, alcohol consumption, physical activity, family history of diabetes, fasting time, use of lipid-lowering medication and aspirin, total energy intake, and BMI. **Abbreviations:** AHEI, alternate healthy eating index; AMED, alternative Mediterranean diet; DASH, dietary approaches to stop hypertension; BMI, body mass index; EDIH, empirical dietary index for hyperinsulinemia; EDIP, empirical dietary empirical dietary inflammatory pattern; HCHS/SOL, Hispanic Community Health Study/Study of Latinos; hPDI, healthy plant-based diet index; HPFS, Health Professionals Follow-up Study; HR, hazard ration; NHS, Nurses’ Health Study; PDI, plant-based diet index; uPDI, unhealthy plant-based diet index; WHI, Women’s Health Initiative.

Approximately 50.0%-65.2% individual metabolites constituting the metabolomic signatures of the 8 dietary indices showed significant associations with incident T2D, and the associations were generally consistent across all cohorts (**Figure 3** and **Figure S11**). For instance, in the combined analysis of all cohorts, 27 out of the 48 metabolites selected for AMED were also significantly associated with T2D risk; notably, 8 metabolites related to a lower AMED score were associated with a higher T2D risk, whereas 6 related to a higher AMED score were significantly associated with a lower risk of T2D. In a sensitivity analysis, the association between all metabolomic signatures and T2D risk persisted when we excluded metabolites from the signatures one at a time, suggesting that the associations between the metabolomic signatures and with T2D risk were likely due to the combined effects of most of the constituting metabolites (**Figure S12**).

We then examined whether, and to which degree the metabolomic signatures mediated the associations between diet and T2D risk. Of the three guideline-based diet, the metabolomic signatures explained 38.2%, 48.5%, and 34.0% of the association between AMED, AHEI, and DASH and T2D risk (FDR < 0.05); a majority of these associations appeared to be mediated by the shared metabolites across the 3 dietary indices (36.5%, 29.3%, and 24.1%, respectively) with minimum contribution from unique metabolites of each diet (**Figure 5** and **Figure S13**). This suggests that the associations between these metabolomic signatures with T2D may be driven by metabolic profiles related to food/nutrient components recommended in all 3 diets. For plant-based diets, their metabolomic signatures explained 20.2%, 17.8% and 5.5% of the associations between PDI, hPDI, and uPDI with T2D risk respectively (FDR < 0.05 except for uPDI). For mechanism-based diets, the metabolomic signatures explained 9.6% and 18.3% of the associations between EDIP and EDIH with T2D risk (FDR < 0.05), with a considerable proportion contributed by both shared metabolites and unique metabolites in each of signatures.

**Figure 5.**
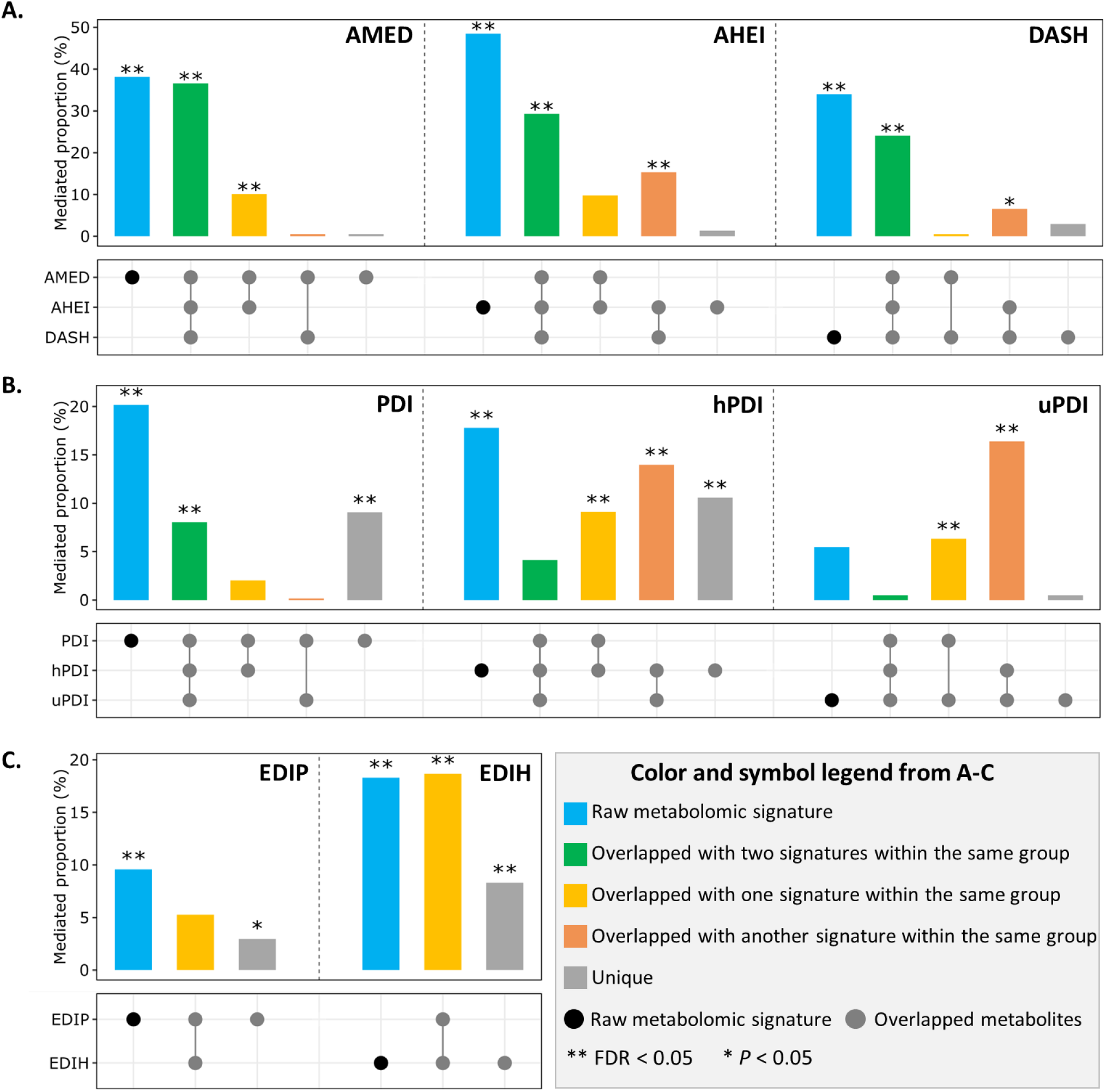
Mediated proportion of shared and distinct metabolites in the relationship of guideline-recommended dietary signatures (A), plant-based dietary signatures (B), and mechanism-driven dietary signatures (C) with incident type 2 diabetes. For this analysis, we combined the results from NHS/HPFS, HCHS/SOL, and WHI**. Abbreviations:** AHEI, alternate healthy eating index; AMED, alternative Mediterranean diet; DASH, dietary approaches to stop hypertension; EDIH, empirical dietary index for hyperinsulinemia; EDIP, empirical dietary empirical dietary inflammatory pattern; HCHS/SOL, Hispanic Community Health Study/Study of Latinos; hPDI, healthy plant-based diet index; NHS, Nurses’ Health Study; PDI, plant-based diet index; uPDI, unhealthy plant-based diet index; WHI, Women’s Health Initiative.

### Genetic determinants of dietary metabolomic signatures

Prior studies showed that genetic variants may affect blood metabolites^36^. We conducted a genome-wide meta-analysis among 13,700 participants from NHS/HPFS, HCHS/SOL, and WHI, to identify genetic determinants of 8 metabolomic signatures of dietary patterns. The SNP-based estimated heritability for metabolomic signatures, on average, ranged between 7.6% (for EDIP) to 19.1% (for hPDI; all *P* < 0.05; **Figure 6A**). We identified a total of 15 unique genetic loci associated with at least one metabolomic signature (loci for each metabolomic signature range from 1 for EDIH, to 9 for DASH; *P*<5×10^-8^ for lead SNPs). Of note, several loci were identified for multiple metabolomic signatures, for example, *FADS1-FADS3* (involved in desaturation and elongation of long-chain fatty acids)^37^ were identified for 7 metabolomic signatures except for EDIH. In addition, *ALMS1*/*ALMS1P1* (involved in synthesis of N-acetylated amino acids)^38^ was identified for 5 metabolomic signatures (namely AMED, DASH, PDI, hPDI, and uPDI) and *TMEM258* (related to lipid metabolism and endoplasmic reticulum stress)^39,40^ for 4 signatures (namely AMED, AHEI, DASH, and EDIP; **Figure 6B** and **Table S10**). Several loci were unique for specific metabolomic signature; for instance, *CERS4* (involved in sphingolipids synthesis)^41^ was identified only for the signature of PDI. In a secondary analysis where we tested gene-diet interaction analysis in associations with metabolomic signature, we identified one locus, *KCNH1* (involved in neurotransmitter release and insulin secretion), whose lead SNP rs148584135 has a significant interaction with AMED score in association with the metabolomic signatures of AMED (*P* = 3.39×10^-8^; **Table S11**).

**Figure 6.**
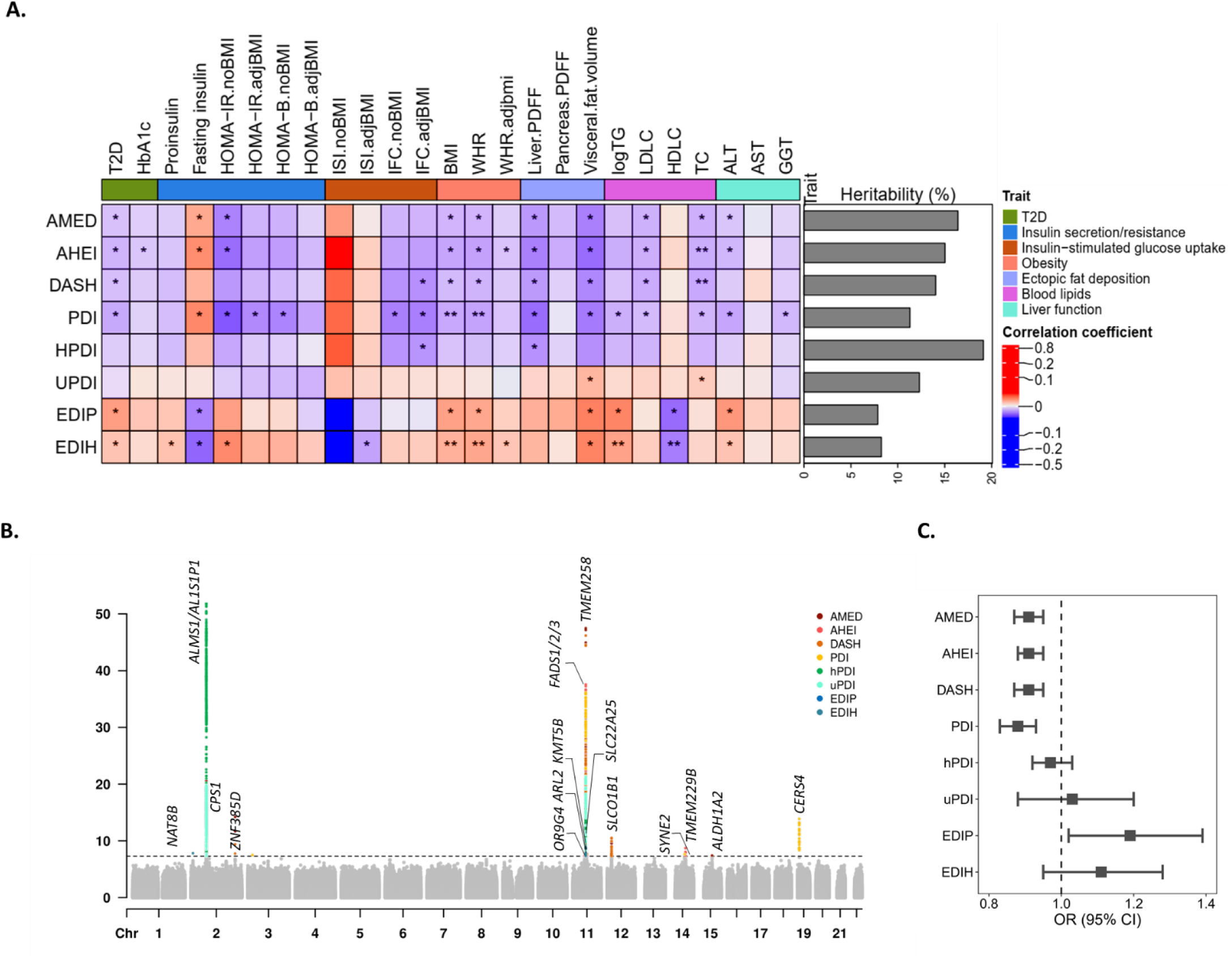
Genetic determinants of dietary metabolomic signatures. **A.** Genetic heritability of 8 metabolomic signatures of dietary patterns (right panel), and genetic correlation between metabolomic signatures and T2D-related traits (left panel). **B.** Genome-wide association meta-analysis of 8 metabolomic signatures of dietary patterns in up to 13,700 individuals from NHS/HPFS, HCHS/SOL, and WHI. The significant *P*-value threshold is 5×10^−8^ (indicated by a grey dash line). **C.** Mendelian randomization analysis between genetically instrumented metabolomic signatures and risk of type 2 diabetes. **Abbreviations:** AHEI, alternate healthy eating index; ALT, alanine transaminase; AMED, alternative Mediterranean diet; AST, aspartate transferase; BMI, body mass index; Chr, chromosome; DASH, dietary approaches to stop hypertension; EDIH, empirical dietary index for hyperinsulinemia; EDIP, empirical dietary empirical dietary inflammatory pattern; GGT, gamma-glutamyl transferase; HbA1c, hemoglobin A1c; HDLC, high-density lipoprotein cholesterol; HOMA-B, homeostatic model assessment for beta-cell function; HOMA-IR, homeostatic model assessment for insulin resistance; hPDI, healthy plant-based diet index; IFC, insulin fold-change; ISI, insulin sensitivity index; LDLC, low-density lipoprotein cholesterol; OR, odds ratio; PDFF, proton density fat fraction; PDI, plant-based diet index; T2D, type 2 diabetes; TC, total cholesterol; TG, triglyceride; uPDI, unhealthy plant-based diet index; WHR, waist-to-hip ratio.

By evaluating whether these mQTLs colocalized with tissue-specific gene expression QTLs across 49 human tissue types, we found that lead SNPs in *FADS1/2*, *TMEM258*, *MYRF, ALMS1/ALMS1P1*—linked to healthy dietary patterns (e.g., AMED, AHEI, DASH, hPDI)—predominantly colocalize with tissues involved in dietary metabolism (adipose, liver, pancreas, and muscle) and endocrine regulation (brain cerebellar hemisphere and hypothalamus), underscoring the pivotal role of dietary metabolism in the pathogenesis of type 2 diabetes (PP.H4.abf > 0.95; **Table S12**).

### Genetic relationship between dietary metabolomic signatures and T2D risk

We further investigated whether genetic variants associated with the 8 metabolomic signatures of dietary patterns are related to T2D development. First, we examined their genetic correlations with a series of clinical traits reflecting various T2D-related pathophysiological pathways: the metabolomic signatures of AMED, AHEI, PDI were genetically correlated with traits reflecting higher insulin sensitivity, less obesity, less visceral and intrahepatic fat deposition, lower low-density lipoprotein (LDL) and total cholesterol, and better liver function (r_g_: -0.14 to -0.25; all *P* < 0.013). The metabolomic signature of DASH was genetically correlated with better post-load insulin response, less obesity and visceral and intrahepatic fat deposition, and lower total cholesterol. In contrast, metabolomic signatures of EDIP and EDIH were genetically correlated with lower insulin secretion and sensitivity, higher obesity and visceral adiposity, lower high-density lipoprotein (HDL) cholesterol, and worse liver function biomarkers (**Figure 6A**).

Metabolomic signatures of 6 out of 8 dietary patterns showed significant genetic correlations with T2D risk (**Figure 6A**). Consistently, in the two-sample MR analysis, we found that the genetically predicted metabolomic signatures of AMED (odds ratio [OR] and 95% CI, 0.91, 0.87-0.95), AHEI (0.91, 0.88-0.95), DASH (0.91, 0.87-0.95), PDI, (0.88, 0.83-0.93), were significantly associated with lower T2D risk; whereas genetically predicted metabolomic signatures of EDIP (1.19, 1.02-1.39) and EDIH (1.11, 0.95-1.28, *P*=0.18) were significantly to marginally associated with increased in T2D risk (**Figure 6C** and **Table S13**), suggesting a potential causal relationship between metabolomic signatures of diets and T2D development.

### Gut microbial factors affecting between-person variation of metabolomic signatures

Gut microbiota play a role in dietary metabolism and have been associated with circulating metabolites^42^. We thus explored the interrelationship between gut microbial taxa and the 8 metabolomic signatures of dietary patterns. In a taxa-wide association analysis, 39 gut microbial species were significantly associated with at least one dietary metabolomic signature(s) (FDR-corrected *P* < 0.05; **Table S14**). For example, the relative abundance of multiple species was identified to be associated with metabolomic signatures of AMED, AHEI, and PDIs enriched with plant-based foods, including *Eubacterium eligens*, *Faecalibacterium prausnitzii, Eubacterium sp. 3_1_31, Butyrivibrio crossotus, Bacteroides cellulosilyticus,* and *Adlercreutzia equolifaciens*^43^ (**Figure S14-S16**). To evaluate how the overall microbial composition was related to the metabolomic signature of each diet, we applied an elastic net regression and calculated a gut microbial score for each of the dietary metabolomic signature – which was a weighted combination of selected microbial species showing strongest association with that signature. We found that across the 3 cohorts included in the analysis (i.e., MLVS, MBS, and HCHS/SOL), the collective microbial composition explained 1.4-7.9% of the variations in metabolomic signatures of guideline-based diets, 1.3-4.7% variation in signatures of plant-based diets, and 1.4-10.6% variation in signatures of mechanism-based diets (**Figure 7A**), suggesting a role of the gut microbiome in potentially affecting diet-related metabolic profiles.

**Figure 7.**
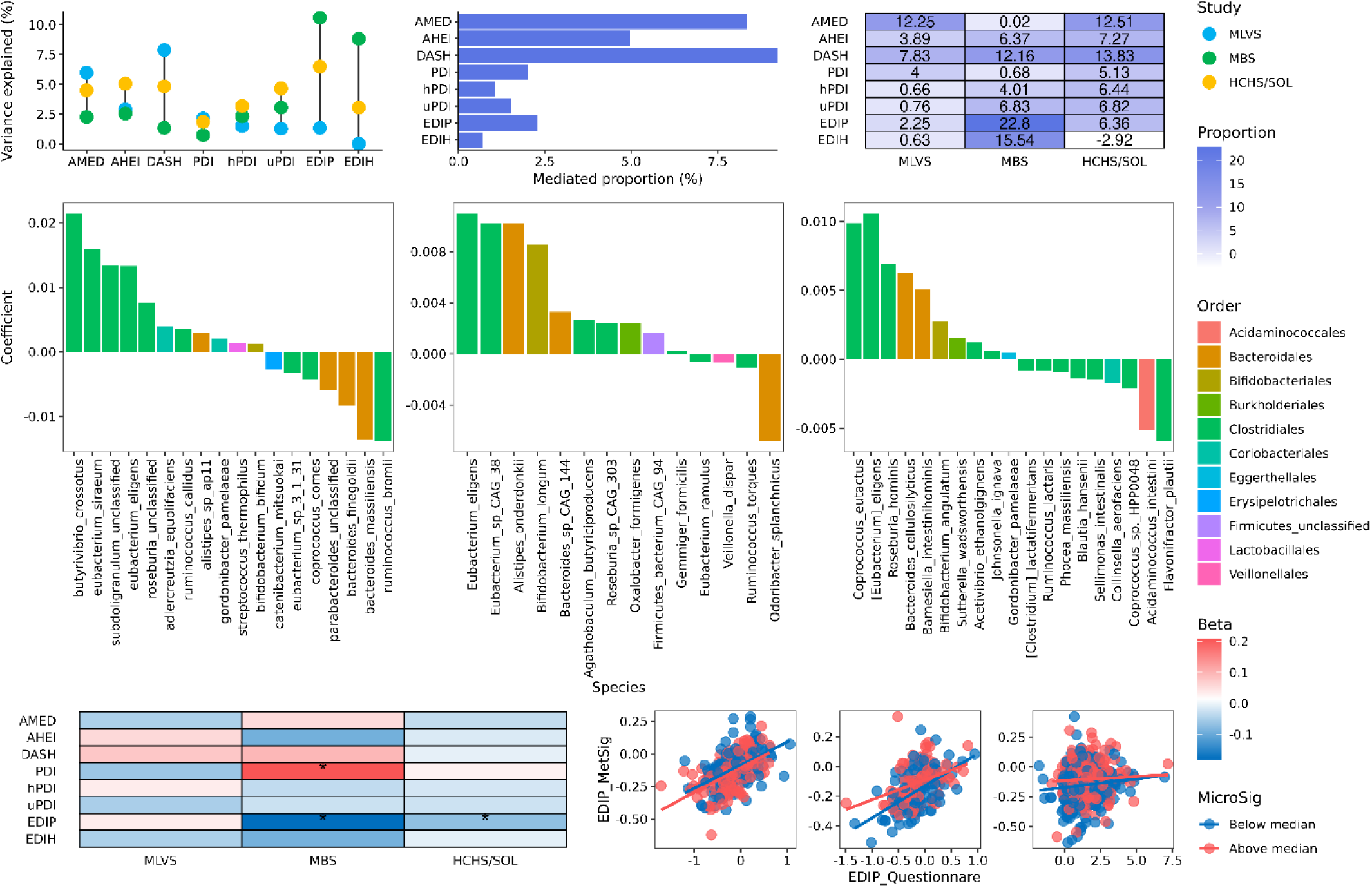
Gut microbial factors affecting between-person variation of metabolomic signatures. **A.** Variance of the metabolomic signatures (MetSig) of dietary patterns explained by gut microbiome in the MLVS, MBS, and HCHS/SOL, respectively. **B.** The mediated proportion of the gut microbiome in the diet-MetSig relationship (left panel: meta mediated association; right panel: mediated association in each study). **C.** Species associated with DASH signature from the elastic net model. **D.** The interaction among dietary patterns, microbial score, and corresponding signatures. **E.** The interaction between EDIP score and gut microbiome on the MetSig for EDIP. **Abbreviations:** AHEI, alternate healthy eating index; AMED, alternative Mediterranean diet; DASH, dietary approaches to stop hypertension; EDIP, empirical dietary empirical dietary inflammatory pattern; EDIH, empirical dietary index for hyperinsulinemia; FFQ, food frequency questionnaire; HCHS/SOL, Hispanic Community Health Study/Study of Latinos; hPDI, healthy plant-based diet index; PDI, plant-based diet index; MBS, Mind Body Study; MLVS, Men’s Lifestyle Validation Study; uPDI, unhealthy plant-based diet index.

We then applied a mediation analysis, to explore the degree to which the overall gut microbial composition explained the associations between dietary pattern indices and their corresponding metabolomic signatures. In the meta-analysis of all 3 cohorts, gut microbial compositions explained a significant proportion of the diet – metabolomic signature associations for all 3 guideline-based diet (i.e., 8.4% for AMED, 5.0% for AHEI, and 9.2% for DASH, all *P* < 0.008; **Figure 7B**). Such a mediating effect was the most consistent across cohorts for DASH (7.83% in MLVS, 12.16% in MBS, and 13.83% in HCHS/SOL); and appears to be largely mediated by species belonging to the *Clostridiales* order, including *Eucbaterium eligens* that appeared in all three cohorts (**Table S15** and **Figure 7B** and **7C**). Microbial compositions mediated 5.13%-6.82% of the associations between PDIs and their metabolomic signatures in HCHS/SOL but less so in other cohorts; and for EDIP and EDIH, their microbial scores mediated 22.80% and 15.54% of the associations between the corresponding dietary indices and metabolomic signatures, respectively, in MBS, but less in other cohorts (**Figure 7B**).

Finally, we explored whether microbial compositions modified the relationship between dietary indices and their circulating metabolomic signatures. We noted a significant interaction between intakes of proinflammatory diet, as quantified by EDIP score from FFQ, and its gut microbial score, in association with the metabolomic signatures of EDIP (*P*_interaction_ = 0.021; **Figure 7D**). Consistently in MBS and HCHS/SOL, the correlation between EDIP and its metabolomic signature was more pronounced among individuals with a lower gut microbial score, compared to those with a higher microbial score (*P*_interaction_ = 0.003 in MBS and *P*_interaction_ = 0.029 in HCHS/SOL; **Figure 7E**). This modifying effect was likely driven by several species including *Megamonas funiformis*, *Clostridium phoceensis*, *Roseburia hominis*, and *Bittarella massiliensis* (**Figure S17**).

## Discussion

Leveraging several large prospective cohort studies with participants of different ethnic backgrounds, our study established and validated unique metabolomic signatures for 8 commonly studied dietary patterns, consisting of both shared and unique metabolites across different diets. Integrated analyses with genetics and metagenomics further showed that, these metabolomic signatures were partially explained by genetic variants related to nutrient and energy metabolism as well as gut microbial compositions, suggesting that these metabolomic signatures also captured variations in metabolic response to diets related to genetics and gut microbiota. Further, the metabolomic signatures were strongly associated with T2D risk independently of self-reported dietary indices, and partially mediated the associations between their corresponding diets and T2D risk. Taken together, we identified metabolomic signatures that assessed the intakes of, and individual metabolic responses to, specific dietary patterns, and were associated withT2D risk.

Some metabolites in biospecimens have been proposed as potentially objective biomarkers to assessed dietary intakes, as complementary to traditional dietary assessment approaches^44^. Several prior studies have identified individual plasma metabolites associated with different dietary patterns (e.g., PDIs, EDIP, EDIH)^10,11^; a recent study^45^ identified metabolomic patterns related to intakes of multiple foods, including fish, dairy, and whole grains, among others. We have previously identified a multi-metabolite signature that assessed the intakes and metabolic responses to a Mediterranean diet, and was associated with CVD risk^12^. The current study, building upon prior progress, is the first to develop multi-metabolite signatures for 8 different yet well-established dietary patterns, by using the elastic net model, the best performing model out of 10 machine learning approaches including decision tree and XGBoost. Many metabolites selected in the signatures for specific dietary patterns are consistent with prior studies; for example, for EDIP and EDIH, our signatures selected caffeine, trigonelline, and N4-acetylcytidine, which have been previously linked to intakes of coffee – a major component in EDIP and EDIH^11,46^. For guideline-based diets, our signatures selected N-acetylornithine due to its strong association with vegetables^47^, 1-methylhistidine for meat^48^, hippuric acid and trigonelline for coffee and tea^49^, and CMPF for fish^50^, with all these associations consistent with previous studies. Our study enhanced the power to assess the overall metabolic profiles in relation to complex dietary patterns by combining the effects of multiple metabolites using machine learning models, and the derived metabolomic signatures showed robust correlation with their corresponding dietary pattern indices in both men and women, and across participants of different racial and ethnic, and age groups. We also utilized metabolites measured by the metabolomic platforms at both the Broad Institute and Metabolon to train the model, therefore enhanced the applicability of our dietary metabolomic signatures in future research.

A unique feature of our study is the comparison of shared and unique metabolites across various dietary patterns. Of the 3 highly correlated guideline-based diets (AMED, AHEI, and DASH) in which recommendations on food groups and nutrients were largely consistent (e.g., less processed foods and more fruits, vegetables, whole grain, nut and legume), their metabolomic signatures shared 32 metabolites whereas only 9%-21% metabolites were unique to each diet. Further, the associations between these guideline-based diets and lower T2D risk were also largely driven by shared (e.g., betaine that may protect against insulin resistance^51^ and acylcarnitine that is related to dysregulated fatty acid oxidation and mitochondrial stress^52^) rather than diet-specific metabolites. In contrast, there were less shared but more diet-specific metabolites when comparing across metabolomic signatures for the 3 plant-based diets and between the EDIP and EDIH. This observation is possibly due to the larger differences and lower correlations in these dietary patterns themselves; for example, hPDI and uPDI weight differently on the included food groups and have only moderate correlation (r= -0.07 to -0.45)^53^; EDIP was empirically developed based on the potentials of food groups in increasing inflammatory biomarkers^54^ whereas EDIH emphasize their potentials in increasing C-peptide^55^. Accordingly, for plant-based and mechanism-based diets, diet-specific metabolites mediated a large proportion of the associations between the dietary intakes and T2D risk. For instance, PE(36:2) and campesterol were unique in EDIP (likely due to their associations with beer and tea), whereas aminoadipic acid and asparagine were specifically selected for EDIH (likely due to their associations with French fries and dairy products); and they are all significantly associated with higher T2D risk in the expected direction. These findings collectively suggested that these metabolomic signatures likely captured the combined metabolic effects of food/nutrient components in their corresponding diets; while generally healthy eating patterns with similar dietary recommendations share similar metabolic pathways in relation to T2D risk, other dietary patterns with specific emphases (e.g., EDIP on dietary proinflammatory potentials) may also associated with T2D risk through more specific metabolic pathways.

The human metabolome reflects the overall combined effects of environmental (e.g., diet) and endogenous factors (e.g., genetics), and the gut microbiome^56^. Through a large-scale GWAS meta-analysis, we identified 15 loci associated with plasma metabolomic signatures of dietary patterns, and found that the SNP-based genetic heritability of the metabolomic signatures were 7.6%-19.1%, reinforcing the notion that host genetic variants may affect the metabolic response to diets. Many of the identified loci are mapped to genes that are involved in biosynthesis and/or catabolism of fatty acids (e.g., *FADS1/2/3* encodes a fatty acid desaturase^37^), phosphatidylcholines (e.g., *TMEM258* is related to protein N-linked glycosylation), sphingolipids (e.g., *CERS4* is involved in ceramide biosynthesis^41^), and bile acids (e.g., *SLCO1B1*^57^). Furthermore, the MR analyses showed a potential causal relationship between metabolomic signatures of the guideline-based diets and PDI with reduced T2D risk, suggesting that genetic variability related to the metabolism of these dietary patterns may play a causal role in T2D development. However, metabolomic signatures of hPDI, uPDI, and EDIH only showed a non-significant MR results but in a direction that was consistent with their epidemiological associations with T2D risk. A possible explanation is the lack of statistical power due to the limited sample sizes of the GWAS of metabolomic signatures. Another potential explanation is that genetic variations related to the metabolism of these diets may play a minimum role in relation to T2D, as compared to the significant associations of these diets themselves with T2D risk. Nonetheless, these data demonstrated that our metabolomic signatures likely captured part of the genetic variations in metabolic responses to various diets, including some genetic pathways that may play a causal role in T2D development.

Another noteworthy finding was the significant relationships between some gut microbial species, predominantly from the *Firmicutes* phylum, and our dietary metabolomic signatures. Previous studies have linked different dietary patterns with changes in microbial compositions^58,59^, and have linked specific microbial species with circulating metabolites^29,30^. Generally in line with prior findings, in our study, metabolomic signatures for AMED, AHEI, DASH, and hPDI are positively associated with the relative abundance of fiber-using bacteria, including *F. prausnitzii*, *B. cellulosilyticus*, and *E. eligens*, which has been related to intakes of healthy plant-based foods^43^. The specific microbial species identified for the dietary metabolomic signatures, and their ability to explain variations in the dietary metabolomic signatures, vary across MLVS (White men aged 71.6 years), MBS (White women aged 60.5 years), and HCHS/SOL (Hispanic/Latino men and women aged 52.6 years), may be somewhat attributed to differences in age, sex, race and ethnicity, and metagenomic sequencing methods. Nonetheless, we observed complex interplay across diets, the gut microbiota, and plasma metabolomic signatures of diets. In particular, the gut microbial compositions (including species e.g., *E. eligens*) partially explained the associations between AMED, AHEI, and DASH and their metabolomic signatures. Moreover, the association between intake of proinflammatory diet and the EDIP metabolomic signatures were also modified by the gut microbial profiles; this may likely because some gut species, e.g., *M. funiformis* (related to higher tea intake and lower inflammatory response^60^), *C. phoceensis* (related to higher fiber intake), *R. hominis* (likely involved in production of TMAO^30^) may play a role in the microbial metabolic processes of specific food components in the EDIP that may affect circulating metabolites in its metabolomic signatures. Collectively, our metabolomic signatures partially reflected inter-individual variation in dietary responses related to gut microbial metabolism.

To the best of our knowledge, this was the first study that systematically identified, validated, and compared the plasma metabolomic signatures for various well-established dietary patterns and their associations with long-term T2D risk, and comprehensively explored the role of genetic variations and the gut microbial compositions in individual metabolic responses to diets. Several limitations warrant discussion. First, we analyzed only metabolomic and dietary data measured at baseline in relation to future T2D risk. Future studies are needed to examine the clinical relevance of these dietary metabolomic signatures with other clinical endpoints, as well as whether changes in diet and the metabolomic signatures over time are related to health using repeated assessments. Second, our study analyzed limited numbers of named metabolites; however, some food-specific metabolites are not measured/annotated by these two metabolomic platforms and some metabolites may not be well measured in our study cohorts (e.g., glucose related metabolites are of low quality in LVS due to delayed sample processing). Therefore, our metabolomic signatures may be inadequate in reflecting some metabolic changes for specific diets, nor other biological changes that are unrelated to the metabolome. Third, due to the observational nature, we cannot make causal inference between dietary intake, the metabolome, and T2D risk. Although we adjusted for multiple confounders and risk factors, residual confounding may exist. Future studies, e.g., dietary feeding studies that comprehensively assess both food metabolome and endogenous metabolites, are needed to validate our signatures and identify novel food-specific biomarkers. Finally, our study population are older adults, and future studies are needed to validate the generalizability of our results in populations of other demographics such as younger adults.

Through integrated analyses of dietary, metabolomic, genetic, metagenomic, as well as clinical data from five prospective cohorts with different ethnic backgrounds, we developed, validated, and compared plasma metabolomic signatures for eight well-established dietary patterns, and showed that these signatures were associated withT2D risk. These metabolomic signatures likely reflect both the intakes of these dietary patterns and individual metabolic responses to diets that are related to host genetic variants and the gut microbiota. These findings may facilitate assessments of dietary intakes and disease risks in clinical and public health settings, thereby informing personalized dietary regimes and more efficient disease prevention.

## Data Availability

All data produced in the present study are available upon reasonable request to the authors.

## Acknowledgements

The authors thank all of the NHS, NHSII, HPFS, HCHS/SOL, and WHI participants for their participation, as well as the staff in these studies for their contribution.

